# Using viral genomics to estimate undetected infections and extent of superspreading events for COVID-19

**DOI:** 10.1101/2020.05.05.20092098

**Authors:** Lucy M. Li, Patrick Ayscue

## Abstract

Asymptomatic infections and limited testing capacity have led to under-reporting of SARS-CoV-2 cases. This has hampered the ability to ascertain true infection numbers, evaluate the effectiveness of surveillance strategies, determine transmission dynamics, and estimate reproductive numbers. Leveraging both viral genomic and time series case data offers methods to estimate these parameters.

Using a Bayesian inference framework to fit a branching process model to viral phylogeny and time series case data, we estimated time-varying reproductive numbers and their variance, the total numbers of infected individuals, the probability of case detection over time, and the estimated time to detection of an outbreak for 12 locations in Europe, China, and the United States.

The median percentage of undetected infections ranged from 13% in New York to 92% in Shanghai, China, with the length of local transmission prior to two cases being detected ranging from 11 days (95% CI: 4-21) in California to 37 days (9-100) in Minnesota. The probability of detection was as low as 1% at the start of local epidemics, increasing as the number of reported cases increased exponentially. The precision of estimates increased with the number of full-length viral genomes in a location. The viral phylogeny was informative of the variance in the reproductive number with the 32% most infectious individuals contributing 80% of total transmission events.

This is the first study that incorporates both the viral genomes and time series case data in the estimation of undetected COVID-19 infections. Our findings suggest the presence of undetected infections broadly and that superspreading events are contributing less to observed dynamics than during the SARS epidemic in 2003. This genomics-informed modeling approach could estimate in near real-time critical surveillance metrics to inform ongoing COVID-19 response efforts.

**Funding:** AWS provided computational credit via the Diagnostic Development Initiative.

## Introduction

SARS-CoV-2 has infected over 3 million people as of May 5, 2020^1^, after first being identified in December, 2019. The rapid expansion of the pandemic, high healthcare and case burdens, and wide observations of mildly symptomatic or asymptomatic infections^2^ have led to continuing uncertainty of the adequacy of public health surveillance systems to effectively estimate the number of cases in a population. Testing capacity remains limited in much of the world, potentially resulting in a large number of infections going undetected.

Estimating the total number of infections is important for several reasons. It provides information on what proportion of the population has been exposed, how many people are still at risk, and the level of community transmission, all critical for determining which public health interventions should be applied and when. Furthermore, estimating the total number of infected individuals helps to evaluate the effectiveness of and identify gaps in surveillance and response efforts. Finally, quantifying the total number of infections provides a more accurate denominator for calculating fatality probability per infection.

The most direct solution to determine the number of infected individuals is to expand population-based testing to identify both symptomatic and asymptomatic cases^3^. However, this would necessitate alternate testing strategies, which has not yet been accomplished outside of relatively limited efforts that are currently impractical to scale in the global response hampered by limited testing capacity. Serological surveys can also inform a post hoc estimate of undetected infections in a population, though this would also require an additional sampling regimen beyond diagnostic testing of active infections and offers limited information on the timing of when infections may have occurred.

Undetected infections can be inferred from gaps in existing data with the use of mathematical models; for example, mobility data and case reports from different geographic locations were used to infer the total number of infections in China^4^. However, the estimate of infection numbers is sensitive to the variance in the offspring distribution (defined as the distribution of secondary infections caused by each infected individual, the mean of which is the reproductive number). That is to say, large outbreaks can occur even if the reproductive number is close to one because of superspreading events^5^. Quantifying the variance around the reproductive number improves the accuracy of estimates of the number of infections and the reproductive number and offers an estimate of the contribution of superspreading events to observed outbreak dynamics.

Viral genomic sequencing is becoming more widespread during viral epidemics^6^ and pathogen genomes can inform estimates of the number of infections as well as the variance around the reproductive number^7^. Integrating the analyses of genomic data into epidemiological mathematical modeling frameworks can jointly estimate reproductive numbers, variance in the reproductive number, and the total number of infections in a population, which are correlated with each other and difficult to estimate using time series case data alone^8^.

Here, we aim to estimate the number of undetected SARS-CoV-2 infections, the time-varying reproductive number and its variance, and the relative contribution of superspreading events to observed outbreak dynamics across time in 12 locations in Asia, Europe, and North America. We use a previously described approach that combines the analyses of phylogenies and time series case data^7^ to arrive at more refined estimates than either source of data can provide independently.

## Methods

### Overview of methods

For each location in our dataset, we fit a branching process model simultaneously to time-series of confirmed cases and to the viral phylogeny of samples from that location. Using a Bayesian inference framework, we estimated the epidemiological parameters of the model as well as latent states. More details are provided in the sections below.

### Data

#### Time series data

We downloaded the time series of confirmed cases for the identified locations from the Johns Hopkins CSSE COVID-19 Github repo^9^ (accessed April 4, 2020). Assuming the dates of disease confirmation were the same dates as symptom onset, we imputed the dates of infection by deducting *U_i_* days from the confirmation date of each case *i*, where *U_i_* was drawn from a Gamma distribution with a mean of 5.5 days and standard deviation of 2.1 days based on estimates of the incubation period of SARS-CoV-2^10^.

#### Viral genomes

We downloaded 1,113 full-length genome sequences from the 12 location from GISAID^11^ (accessed April 4, 2020): 1) California, US, 2) Minnesota, US, 3) New York, US, 4) Washington, US, 5) Guangdong, China, 6) Hong Kong, China, 7) Hubei, China, 8) Shanghai, China, 9) Iceland, 10) Italy, 11) Japan, and 12) United Kingdom. Inclusion criterion for geographic locations was the availability of ≥ 50 full-length SARS-CoV-2 genomes from countries or ≥ 30 from first-level administrative divisions as of April 4, 2020 from GISAID database. The following locations also satisfied this criterion but were excluded due to multiple circulating lineages:

Austria, Belgium, and Ontario, Canada. We also excluded the following locations because the model did not converge within the timeframe of the analysis: France, Netherlands, and Switzerland. We excluded sequences with more than 10% ambiguous sites. The location of each sequence was assigned according to the annotation on GISAID or the source of exposure, if available. For each of the 12 locations, we aligned the sequences against each other using MAFFT v7.455^12^ with default settings.

We used the ModelFinder^13^ program within IQ-TREE v1.6.12^6^ to identify the best-fit substitution model for each dataset according to the Bayesian Information Criteria, and then used the selected model to infer the maximum likelihood phylogeny in IQ-TREE v1.6.12 allowing for multifurcations. The command line options to IQ-TREE v1.6.12 were *-bnni -czb*.

Assuming a molecular clock rate of 8 x 10^-4^ with a standard deviation of 5 x 10^-4^ substitutions per site per year^14^, we used TreeTime to estimate the dates of branching events in the phylogeny and re-rooted the phylogeny to maximize the correlation coefficient of the root-to-tip plot. The command-line options for treetime were --reroot least-squares --clock-filter 3 --tip-slack 3 --confidence --clock-rate 0.0008 --clock-std-dev 0.0005. The resulting time trees are provided in Supplementary Data 1.

To minimize the effects of multiple introductions and co-circulating lineages on the estimation of infection numbers, we subsampled the sequences to the dominant lineages. The excluded samples are listed in Supplementary Table 1.

#### Analysis time periods

For each dataset, we estimated the number of new infections starting 10 days before the first reported case. The end dates were the most recent collection dates of sequences available on GISAID on April 4, 2020. As such, the end dates of analysis were different across the 12 geographic locations.

### Model inference

We fit a stochastic branching process model to both the time-series data and viral phylogeny, seeded by *N_0_* number of infections. The number of secondary infections *v_i_*, caused by each individual *i* infected during time step *t* was drawn from a negative binomial distribution with mean *Rt* and coefficient of variance *ψ*. The times *τ_j_* of onward transmission to individual *j* relative to the infection time of the transmitter were drawn from a Weibull distribution with mean of 5.0 days and standard deviation of 1.9 days^10^. The probability of detecting an infection during time step *t* is given by *ρ_t_*. The latent variable *x_t_* tracks the number of new infections over time.

We jointly estimated the latent variable *x_t_* jn addition to the following parameters for each location using Bayesian inference: *R_t_*, *ψ, ρ_t_*, *N*_0_. The shape *α* and scale *s* parameters of the

Weibull distribution are fixed to give the mean and standard deviation listed above. The list of parameters, their prior distributions or fixed values are listed in Supplementary Table 2.

Using the Bayesian particle Markov chain Monte Carlo (PMCMC) with Metropolis-Hastings sampling algorithm described in Li et al.^8^, we fit the branching process model to the time series *y*_1:_*_T_* and time-resolved viral phylogeny *z*_1:_*_T_* for each of the 12 locations. We randomly generated 100 sets of initial parameter values from the prior distributions, ran the PMCMC for 1000 iterations, and then picked the parameter combination for each dataset with the highest posterior probability to start the final PMCMC inference. We ran the final PMCMC inference for up to 1 million iterations sampling every 100 iterations, stopping the runs earlier if at least 300 effective samples were obtained. We checked convergence visually and by calculating the effective sample size. In all PMCMC runs, we used 5,000 particles to simulate the epidemics and iteratively calculated the likelihood every 8 time steps, where each time step was 0.25 days.

During each iteration of the PMCMC algorithm, a new value for one of the parameters in *θ* is drawn from a truncated normal distribution centered around the last accepted parameter value. The bounds of the truncation are listed in Supplementary Table 2. The new set of parameter values *θ′* are accepted with probability 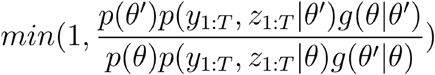 where *p*(*θ*) is the prior probability of parameters *θ* and *g{θ|θ′*) is the density function of proposing *θ* given *θ′*, and vice versa.

We concurrently estimated the likelihood 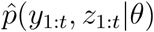 of parameters *θ* = (*R_t_, ψ, ρ_t_, N*_0_) and obtained a sample of the latent epidemic trajectory *x*_1:_*_T_* from the density *p*(*x*_1_*_:T_|y*_1:_*_T_, z1:T*). We iteratively estimated the likelihood 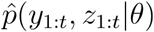 of parameters *θ* at each time step *t =* 1,…,*T* by simulating *N* epidemic trajectories 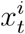 for *i* = 1,…,*N* using the stochastic branching process model, calculating 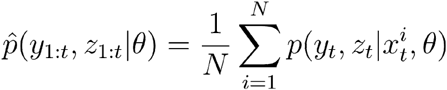, and sampling *N* particles in the next step according to a multinomial distribution with probabilities proportional to 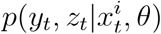. The epidemic trajectory *x_t_* defines the number of newly infected people at time step *t*. Detailed explanation of the PMCMC algorithm has been previously described^15,16^, as well as its application for fitting epidemiological models to both time series data and phylogenies^8,17^.

The probability 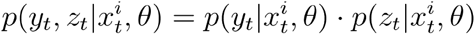 defined as follows. Let 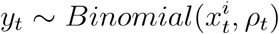, then 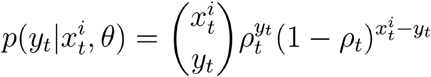 where *ρ_t_* is the probability of detecting an infection during time step *t*.

For a given epidemic trajectory 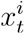, the coalescent (branching) rate *λ_t_* is given by 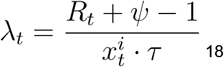^18^. Let 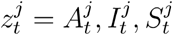, for *j* = 1,…,*M_t_*, where *M_t_* is the number of intervals within the phylogeny during time step *t*, 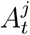 is the number of lineages during that interval, 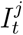 is an indicator variable that equals 1 when the interval starts with a branching event and 0 otherwise, and *S_t_* is the time duration of the interval. The number of intervals during a time step is one more than the number of events during that time step, where an event is defined as a branching or sampling event. The probability 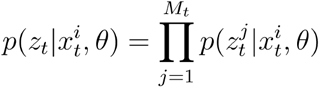, where

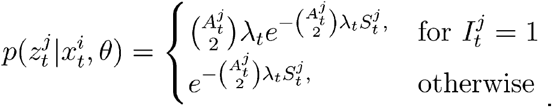

Both the reproductive numbers *R_t_* and reporting rates *ρ_t_* were estimated for each one-week period starting from the first reported case. The *R_t_* and *ρ_t_* parameters before the first reported case were assumed to be the same as *R_t_* and *ρ_t_* values during the first week of the reported case time series.

### Code availability

Code to carry out data cleaning and visualization is available at https://github.com/czbiohub/EpiGen-COVID19 and the code to run the model is available at https://github.com/lucymli/EpiGenMCMC.

### Role of funding source

The computational aspects of this work were supported by the AWS Diagnostic Development Initiative via computational credit.

## Results

### The proportion of unreported infections

We jointly estimated the parameters of the branching process model and the epidemic trajectories, i.e. the number of new infections over time, using the Bayesian inference algorithm described in Methods. Unless defined otherwise, the numbers provided are the median estimates with the 95% highest posterior density interval noted in the parentheses.

Defining the number of undetected infections as the estimated number of infections above the number of infections that were reported in the time series, we found that 30% (95% HPD: 13%-50%) of infections across all locations across all time points were not detected in time series of confirmed cases. This median estimate varied from 13% and 92% across locations (Table 1; Figure 1).

**Table 1.** The estimated numbers of undetected SARS-CoV2 infections across the 12 locations in the study during the analysis periods, and the number of undetected infections before the first confirmed case. The ‘Undetected infections’ columns are rounded to 2 significant figures to reflect uncertainty in estimates. The 95% highest posterior density intervals are indicated in parentheses beneath median estimates. The dates are in the format ‘year-month-day’.

| Location | Reported cases<br>(during analysis<br>period) | Undetected infections<br>(during analysis<br>period) | Undetected infections (%)<br>(during analysis period) | Undetected<br>infections (before<br>first reported case) | Analysis dates | Date of first<br>reported<br>case |
| --- | --- | --- | --- | --- | --- | --- |
| California,<br>US | 2,690 | 1,600<br>(83 - 9,400) | 37.2%<br>(3.0% - 77.7%) | 31<br>(0 - 320) | 2019-12-30 to<br>2020-03-17 | 2020-01-21 |
| Minnesota,<br>US | 108 | 340<br>(38 - 1,400) | 75.6%<br>(26.0% - 92.9%) | 210<br>(27 - 850) | 2020-01-28 to<br>2020-03-12 | 2020-03-02 |
| New York,<br>US | 21,740 | 3,200<br>(370 - 13,000) | 12.8%<br>(1.6% - 36.8%) | 160<br>(0 - 1,000) | 2020-01-22 to<br>2020-03-16 | 2020-02-25 |
| Washington,<br>US | 4,563 | 7,300<br>(3,800 - 13,000) | 61.4%<br>(45.5% - 73.3%) | 1,500<br>(970 - 2,100) | 2020-01-15 to<br>2020-03-23 | 2020-02-18 |
| Guangdong,<br>China | 1,298 | 890<br>(100 - 2,900) | 40.6%<br>(7.4% - 68.8%) | 260<br>(40 - 790) | 2019-12-07 to<br>2020-02-09 | 2020-01-10 |
| Hong Kong,<br>China | 96 | 340<br>(16 - 1,500) | 78.0%<br>(14.6% - 93.9%) | 80<br>(3 - 310) | 2019-12-13 to<br>2020-02-23 | 2020-01-16 |
| Hubei,<br>China | 29,992 | 10,000<br>(4,100 - 35,000) | 25.8%<br>(12.1% - 53.7%) | 280<br>(61 - 740) | 2019-11-02 to<br>2020-02-02 | 2019-12-06 |
| Shanghai,<br>China | 323 | 3,900<br>(2,200 - 6,500) | 92.3%<br>(87.4% - 95.3%) | 1,900<br>(1,200 - 2,900) | 2019-12-09 to<br>2020-02-08 | 2020-01-12 |
| Iceland | 673 | 1,100<br>(310 - 2,400) | 61.6%<br>(31.4% - 78.1%) | 370<br>(120 - 760) | 2020-01-21 to<br>2020-03-17 | 2020-02-24 |
| Italy | 64,272 | 27,000<br>(8,800 - 56,000) | 29.6%<br>(12.0% - 46.7%) | 420<br>(14 - 2,200) | 2019-12-23 to<br>2020-03-16 | 2020-01-26 |
| Japan | 247 | 940<br>(160 - 3,000) | 79.2%<br>(39.6% - 92.4%) | 150<br>(23 - 430) | 2019-12-14 to<br>2020-02-23 | 2020-01-17 |
| United<br>Kingdom | 23,216 | 6,100<br>(2,600 - 12,000) | 20.8%<br>(10.0% - 34.1%) | 820<br>(370 - 1,600) | 2019-12-21 to<br>2020-03-24 | 2020-01-24 |

**Figure 1.**
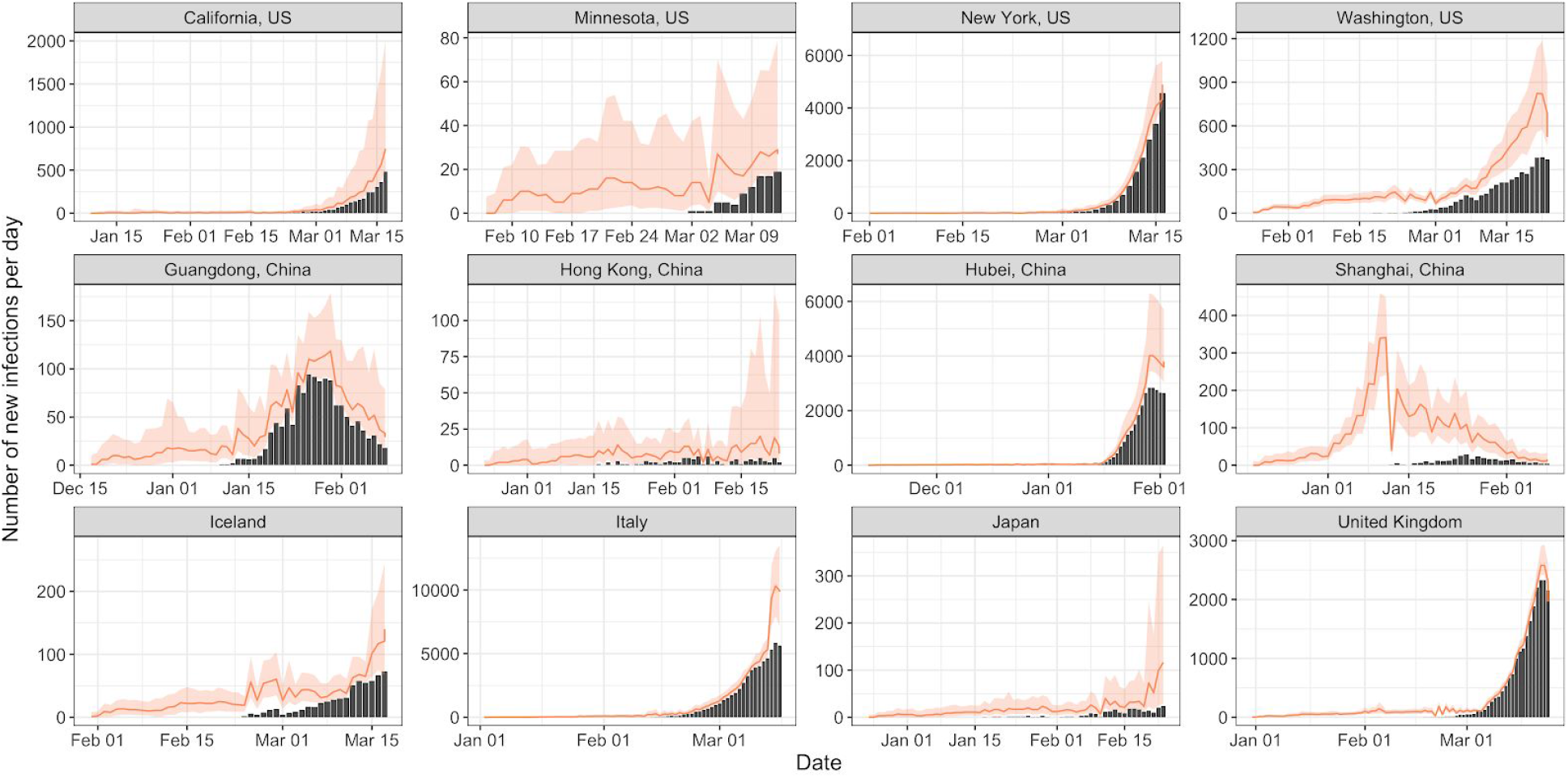
The estimated number of new infections over time across the 12 locations (red dots represent the median estimates on those days, and the red lines indicate the 95% highest posterior density intervals on those days). The bars are the number of reported confirmed cases by imputed date of infection.

We found that between January 10 and 23 there were 4,900 (95% HPD: 4,100–7,600) total new infections in Hubei province, which was lower than the previous estimate for Wuhan city (capital of Hubei) of 13,118 (95% CI: 2,974–23,435)^4^, though we were able to obtain more precise estimates and the credible intervals overlapped. Similarly, our median estimate of 2,100 (1,200–3,900) was lower but the credible interval overlapped with the 4,000 (1,700–7,800) total new infections estimated for Wuhan based on traveler data up to January 18^19^. Finally, 20,767 (9,528–38,421) infections were estimated for Wuhan up to January 29 based on data from an evacuation flight to Japan^20^, which is on par with our finding of 21,000 (95% HPD: 18,000–33,000) during the same period.

The number of undetected infections in California and New York state was unexpectedly small given reports of delayed reporting^21^. To evaluate if oversampling of viral genomes in New York, Nassau, and San Francisco Bay Area counties may contribute to biased state-level observations, we re-calculated the total numbers of undetected infections for California and New York using only confirmed cases from those counties. The estimate of proportion of infections that were undetected increased from 37.2% (3.0% - 77.7%) to 78.9% (67.9% - 92.0%) for California, and from 12.8% (1.6% - 36.8%) to 40.0% (32.0% - 58.7%) for New York. Further analysis of just time series data from these counties may further support these adjusted proportions.

The credible intervals around the proportion of infections that were undetected in each location was inversely correlated with the number of sequences available for that location (Supplementary Figure 2). To quantify the association, we fit a beta regression model of the form *logit*(*Y*) = *a − b · log*(*X*) _w_here *Y* is the size of the uncertainty interval around the proportion of undetected infections (Table 1, column 4) and *X* is the number of sequences used for analysis. The best fit parameters were *a* =2.08 (0.59-3.57) and *b*=0.54 (0.20-0.89), which meant that increasing the number of sequences from 10 to 100 decreased the uncertainty interval size by 43% from 70% to 40%, and increasing the number of sequences from 100 to 500 further reduced the uncertainty interval size by 46% from 40% to 21%.

Across all locations, the lowest probability of detection occurred within the preliminary phase of the epidemic (Figure 2). This is in line with another modeling study that found increasing rates of detection in the US across time^22^. The estimate of the overall probability of detection based on pandemic data from China February 7, 2020 ranged from 7.1% to 47.6% (95% confidence interval: 31.6-86.9%)^23–25^, which are not directly comparable to the estimates in this study as they used data from all of China rather than from a specific province.

**Figure 2.**
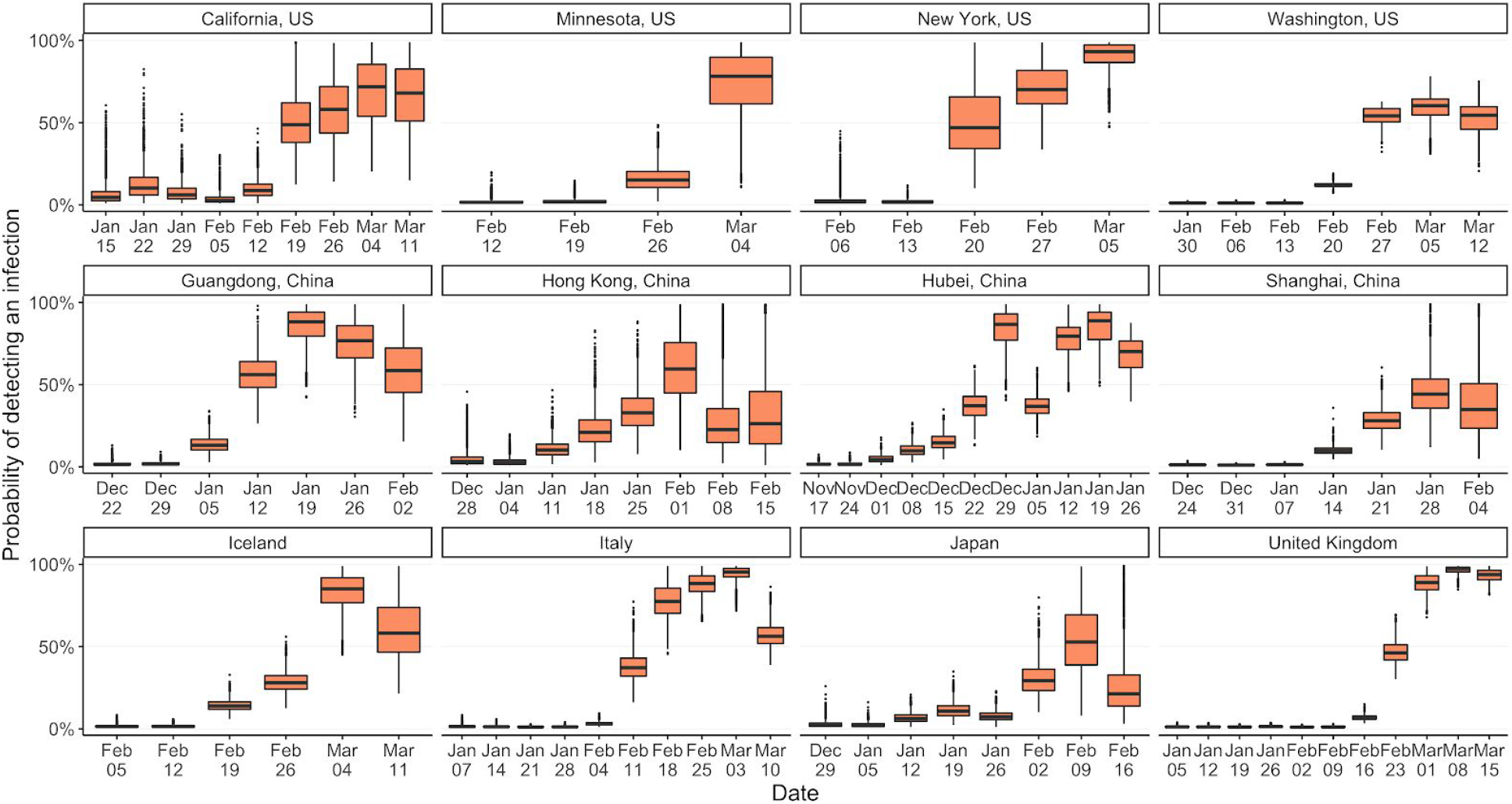
The probability of detecting a case *P_t_* during each week of the analysis period.

### Heterogeneity in reproductive numbers

The median reproductive numbers *R_t_* during each week was above one in 70% of the weeks analyzed across the 12 locations (Figure 3). This percentage ranged from 43% in Shanghai to 90% in Italy.

**Figure 3.**
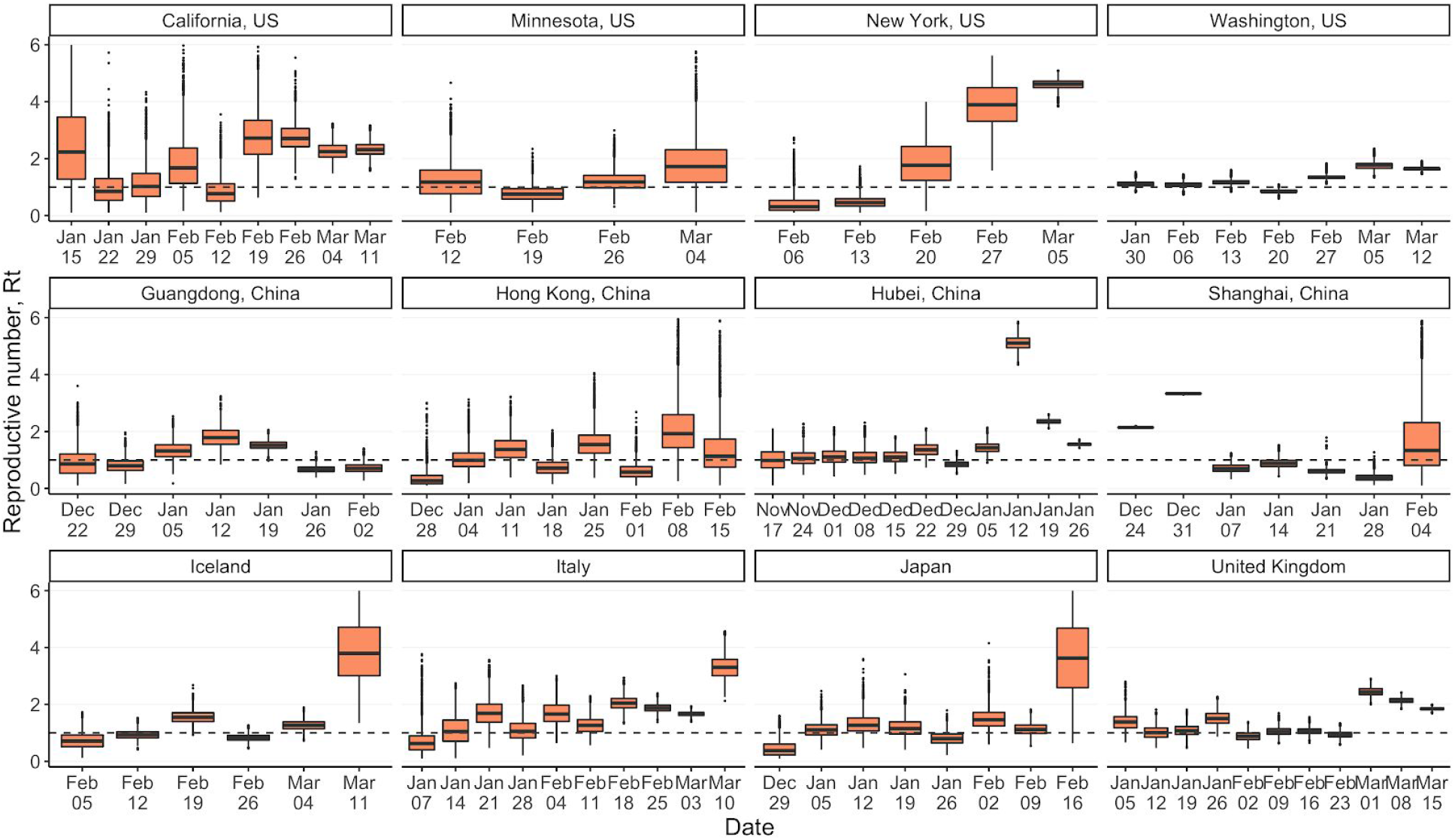
The reproductive number *R_t_* over time across the 12 locations. The dates indicate the start of the week for which the *R_t_* was estimated.

The overall reproductive number across all locations and time periods was 1.20 (0.12-3.34), with the initial reproductive number in each location ranging from 0.27 (0.10-0.93) in Hong Kong to 2.23 (0.20-5.32) in California.

To quantify the variance in the reproductive numbers and evaluate the contribution of superspreading events to epidemic dynamics, we computed the smallest number of individuals that could contribute to 80% of infections during each week (Figure 4). The smaller the number, the larger the variance and the more likely that there are superspreading events such as those observed during the SARS outbreak of 2003. Across all locations and time periods, the top 33% (2%-47%) of infected individuals contributed to 80% of onward infections. This was higher than the 16% and 10% estimated for the SARS outbreak in Singapore and Beijing, respectively^5^. The numbers were calculated from the estimated *R_t_* (Figure 3) and coefficient of variation *ψ* values (see Supplementary Figure 1).

**Figure 4.**
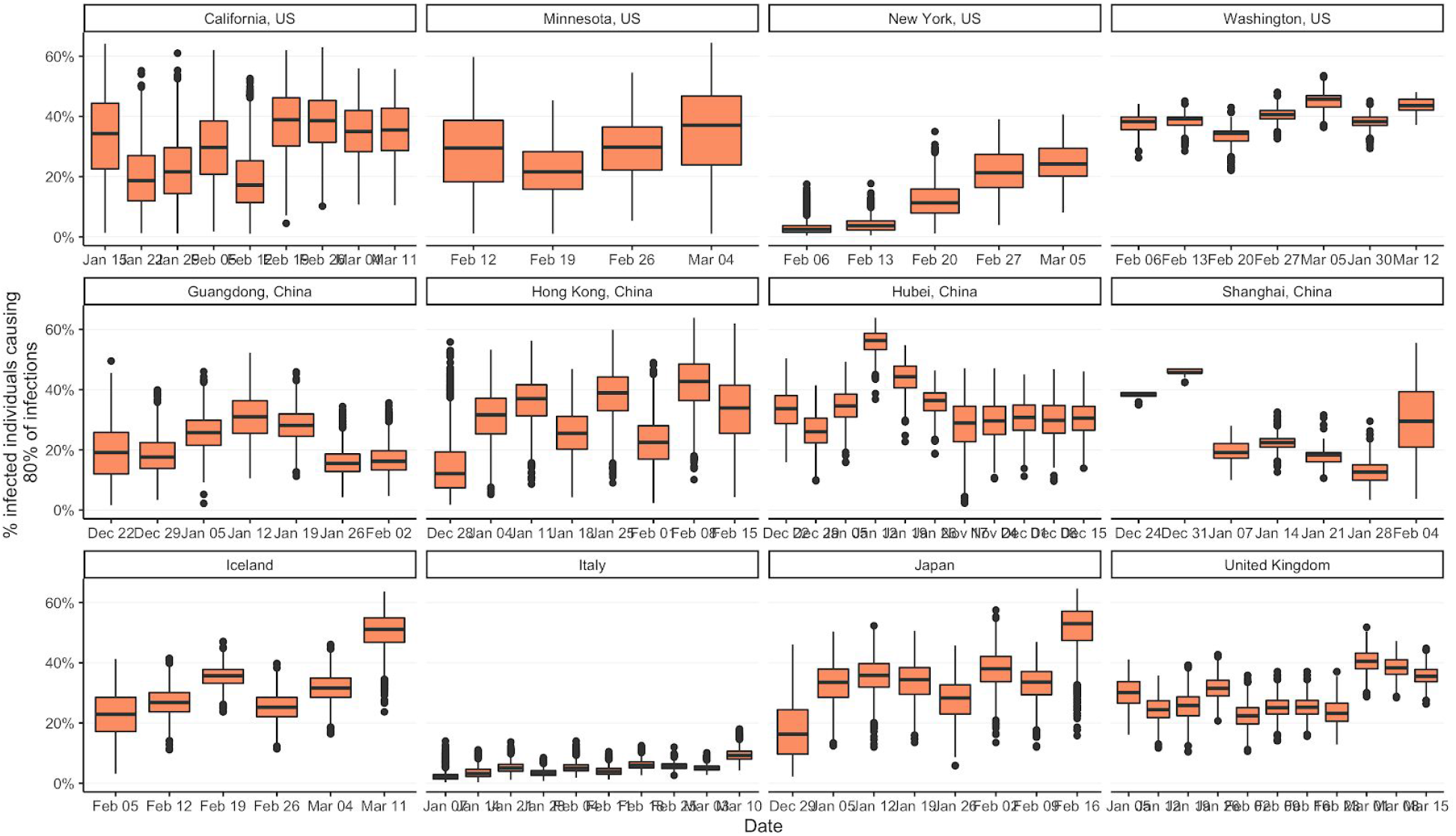
The top percentage of individuals causing 80% of infections, given the negative binomial offspring distribution during each week with mean *R_t_* and coefficient of infection *ψ*. The lower the percentage, the larger the variance around reproductive number.

Another common parameterization with negative binomial distributions of the reproductive number is the dispersion parameter *k* = *R_t_/*(*ψ* − 1). The smaller the value of *k*, the greater the variance. The estimated values of *k* are 0.38 (0.01-3.46) for reproductive numbers between 1 and 2, and 0.69 (0.02-6.35) for reproductive numbers between 2 and 3.

### Time until a local outbreak is detected

Given the probabilities of detection *ρ_t_*, the reproductive number *R_t_*, and the coefficient of variation *ψ* at the start of the analysis in each location, we estimated the length of time that would pass before local spread of COVID-19 was detected. We simulated from the branching process model using the highest posterior probability parameter combination in each location to determine the length of time it would take for 2 infections to be detected in a new location, assuming that local transmission would be recognized after 2 confirmed cases.

If outbreaks occurred in similar settings to the 12 locations in terms of initial reproductive numbers and detection probabilities, we estimate 11 to 37 days would elapse before 2 infections were detected, by which point there would be 4 to 45 total infections in the local population (Table 2).

**Table 2.** The expected number of days before 2 infections were detected in a new location. This was based on simulations from the branching process model using initial parameter values estimated for the 12 locations in this study. The last column denotes the expected number of total infections by that date. We used maximum posterior density *R_t_*, *ρ_t_*, and *ψ* values to generate simulations from which we calculated the values in the last two columns. The 95% confidence intervals are shown in parentheses.

| Location | $R_t$ | $\rho_t$ | $\psi$ | Days to notice | Total infected |
| --- | --- | --- | --- | --- | --- |
| Italy | 0.3 | 1.3% | 39.3 | 22 (6-28) | 45 (4-232) |
| New York, US | 0.3 | 1.6% | 11.9 | 20 (6-50) | 28 (3-104) |
| Japan | 0.6 | 1.2% | 1.3 | 32 (9-79) | 4 (2-13) |
| Hong Kong, China | 0.6 | 1.0% | 2.1 | 31 (8-61) | 8 (2-24) |
| Minnesota, US | 0.9 | 1.1% | 3.8 | 37 (9-100) | 11 (2-48) |
| Iceland | 1 | 1.1% | 2.2 | 35 (8-95) | 9 (2-29) |
| Washington, US | 1.2 | 1.0% | 1.5 | 35 (9-91) | 9 (3-26) |
| Hubei, China | 1.2 | 1.6% | 1.7 | 25 (7-65) | 8 (3-28) |
| United Kingdom | 1.4 | 1.2% | 2.8 | 28 (8-64) | 13 (3-42) |
| Guangdong, China | 1.6 | 1.4% | 3 | 23 (7-51) | 15 (4-48) |
| Shanghai, China | 2.1 | 1.0% | 2.8 | 22 (7-43) | 21 (4-87) |
| California, US | 4.3 | 2.2% | 3.3 | 11 (4-21) | 24 (5-73) |

## Discussion

The identification of asymptomatic individuals presents a particular challenge to COVID-19 surveillance and outbreak response. Serosurveys will help inform estimates of the proportion of the population that has been exposed cumulatively but have limited utility in evaluating time-varying case ascertainment. Our findings using viral genomic data suggest that local introductions of SARS-CoV-2 resulted in sustained transmission occurring for days or weeks before infections were detected in every location evaluated. The probability of detection has been dynamic across time as diagnostic testing capacity and response strategies have been operationalized. These data further suggest that aggressive response following identification of an index case may be warranted to identify cryptic chains of transmission. The approach we deploy here may also be useful for monitoring the performance of COVID-19 surveillance efforts, including if the rate of undetected infections will continue a decreasing trend as new diagnostic assays and testing strategies are developed throughout the course of the pandemic.

The inclusion of the genomic data in the modeling of the spread of COVID-19 enabled more precise estimates of the number of infections, and the precision increased with more genomic sequences available. Additional viral sequences should further improve epidemiological estimates. The data were downloaded for the current analysis on April 4 and as of April 30 the total number of full-length viral genomes has increased from 3,001 to 4,148. Furthermore, given the increasing availability of serological tests, integrating those results within a modeling framework can further constrain the set of plausible epidemic trajectories.

The viral phylogeny was also informative of the variance around the reproductive numbers. The level of variation estimated here was on par with what has been estimated for pandemic influenza^21^ but not as variable as observed in SARS or MERS^4,22^, suggesting a smaller contribution of COVID-19 superspreading events to observed outbreak dynamics and that a large proportion of the population can contribute to onward infections. It is worth noting, however, that variance was significantly larger in Italy. This could be indicative of nosocomial settings amplifying transmission, though more epidemiological studies are warranted to determine the source of relative low variance in the Italian epidemic.

Another modeling study^26^ using outbreak sizes in different countries estimated a larger variance in the reproductive number, with dispersion parameter *k* estimated to be 0.10 (0.05-0.20) for reproductive numbers between 2 and 3, in contrast to 0.69 (0.02-6.35) estimated in the current study. The dependence on the final outbreak size distribution to estimate *k* could explain the difference in estimates, as there are different delays in returning testing results in different countries and final outbreak sizes are still not available for most regions as the pandemic is ongoing. The larger credible interval around estimate in the present study reflects the wide range of location- and time-specific estimates of reproductive numbers.

One of the limitations of this study was that we did not take into account time-varying delays in testing during an individual’s course of infection, so the time series data could be offset from the genetic data by a number of days. This is a reasonable assumption in locations with consistent testing capacity, however these delays likely changed across the course of the epidemic with changing testing regimens and burdens on the healthcare and public health systems. More data on testing capacity over time in different locations could support refined parameterization of the delay distribution over the course of the pandemic.

Using genomic data to infer infected numbers generally is not as sensitive to sampling schema as serological surveys, though we do see signals in the data that opportunistic genomic sampling may be over-representing subpopulations and biasing estimates for a subset of locations. This could explain the seemingly high detection rates in California, New York, and the United Kingdom, where multiple smaller outbreaks are occurring within the country or state, but genomic data were only generated from a subset of those locations, biasing results towards underestimation of the total number of cases at the state or country level. Conversely, the estimated infection numbers are likely biased upwards near the root of the phylogeny due to multiple introductions into each location^27^. This would unlikely impact the overall detection rate as those early infections only accounted for a small percentage of the total number of infections. Future work will aim to generate more representative viral genomic sampling within evaluated regions.

The relatively high number of undetected infections in Iceland was unexpected given the extensive efforts to test the population. However, population screening only began on March 13, towards the end of the analysis period^20^. Re-running the analysis to accommodate the results of the widespread testing should provide estimates of how much the population screening increased detection rates.

Spatial and age structures were not explicitly parameterized by the branching process model, although the variance of the reproductive number does capture some of the heterogeneity in transmission due to population structure. More explicit parameterization of spatial structure can facilitate more spatially granular estimates of epidemiological parameters, such as with mobility data. However, incorporating spatial structure in the coalescent model can reduce the identifiability of variance of the reproductive numbers^27^.

In addition to improving retrospective parameter estimates, the estimates of detection probabilities and reproductive numbers can be used to parameterize predictive transmission models to project future infection numbers. Data on testing rates could also improve calibration of estimated detection probabilities.

Given the changes in undetected infections and reproductive numbers from week to week during the dynamic pandemic response, repeating the analysis outlined in this paper at multiple time points can provide near real-time estimates of the reported parameters. Simulations of genomic data from the model presented here could also help optimize sampling strategies to estimate total infection numbers, particularly for regions which have not yet experienced exponential growth of COVID-19 cases.

The genomics of SARS-CoV-2 has played a crucial role in tracking COVID-19 spread globally as well as viral evolution. We demonstrated that the viral phylogeny is also informative of infection numbers, reproductive number and its variance, and detection performance of COVID-19 surveillance systems, critical to informing effective public health response strategies. Continued efforts to share full-length viral genomes via open-source databases will enable additional tools in the effort to track and respond to the COVID-19 pandemic.

## Data Availability

Data are provided as supplementary files.

## Acknowledgements

The computational aspects of this study were supported by the AWS Diagnostic Development Initiative. We acknowledge the helpful feedback on the manuscript from Jack Kamm, Josh Batson, and Joseph Bradley.

## Authors’ contributions

Lucy M. Li: conceptualization, analysis, data curation, visualization, interpretation of results, writing. Patrick Ayscue: interpretation of results, writing.

## Supplementary materials

Supplementary Table 1. List of GISAID sequences included in the analysis and associated metadata.

Supplementary Table 2. Prior distributions of model parameters that were estimated and parameter values that were fixed based on literature.

Supplementary Table 3. The median estimated negative binomial dispersion parameter 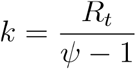 and coefficient of variation *ψ* for each range of estimated *R_t_* values. The 95% highest posterior density intervals are shown in parentheses.

Supplementary Figure 1. Coefficient of variation *ψ* of the reproductive number *R_t_*, across the 12 locations.

Supplementary Figure 2. The size of the uncertainty interval around the undetected infection estimates (Table 1 column 4) decreased with the number of sequences available for analysis at that location. The blue line is best-fit beta regression model predictions, and the grey shaded region is the uncertainty interval at the 2.5% and 97.5% quantiles.

Supplementary Data 1. Phylogenies for the 12 locations in nexus format.

Supplementary Data 2. Time series of new infections in the 12 locations.

## Notes

### Competing Interest Statement

The authors have declared no competing interest.

